# Public Perceptions and Discussions of Premium Cigars on Reddit

**DOI:** 10.1101/2023.06.22.23291751

**Authors:** Zidian Xie, Sarah Lee, Emily Xu, Dongmei Li

## Abstract

**Introduction:** While premium cigars have similar addictive, toxic, and carcinogenic constituents as other cigars and cigarettes, about 1% of the US adults reported premium cigar use from 2010 to 2019. This study aimed to understand public perceptions and discussions of premium cigars on Reddit, one of the most popular social media platforms.

**Methods:** Using keywords such as “premium cigar”, we extracted 2,238 Reddit posts from Reddit Archive between July 2019 and June 2021. Among them, 1,626 posts were related to premium cigars. By employing the inductive approach, we manually coded each Reddit post on premium cigars to understand public perceptions and discussions of premium cigars by summarizing them into different topics and subtopics.

**Results:** Longitudinal analysis showed that the number of Reddit posts on premium cigars increased since June 2020. Content analysis showed that among Reddit posts related to premium cigars, the most popular topic is “Information sharing” (75.72%), in which Reddit users shared their perceptions about premium cigars, asked for advice, and provided some recommendations about premium cigars. Over one-quarter of posts (27.17%) are sharing user experiences of premium cigars (such as taste). Nearly one-fifth (18.99%) of posts are discussing the affordability of premium cigars. In addition, 7.87% of posts are discussing legal/policy issues related to premium cigars, and 6.82% of posts are related to the health risks of premium cigars compared to cigarettes.

**Conclusions:** Public perceptions including misperceptions, user experiences, and affordability related to premium cigars have been actively discussed on Reddit.

**Implications:** With the increasing trend of premium cigar use, it is necessary to understand how premium cigars are perceived by the public and why they are becoming more popular. This study provides the first evidence on public perceptions and discussions of premium cigars on social media, which could provide useful information on future regulatory policies that aim to prevent the prevalence of premium cigars to protect public health.

## INTRODUCTION

As the leading cause of preventable death and diseases (such as respiratory and heart diseases), tobacco use is responsible for the death of 8 million people annually on a global scale ^1^. While the consumption of cigarettes has decreased, the use of cigars has been increasing in recent years ^2^. Based on Nielsen data, the cigar sales in the US have increased from $2.47 billion in 2009 to $3.27 billion in 2020 ^3^. Cigars have three main types, including large cigars, cigarillos, and little cigars. Based on the recent definition from the National Academies of Sciences, Engineering and Medicine ^4^, a premium cigar is handmade, composed of at least 50% natural long-leaf filler tobacco, large size, wrapped with leaf tobacco, no characterizing flavor, and without filters or tips. The 2012-2013 National Adult Tobacco Survey showed that about 19.9% of U.S. adults who smoke cigars smoked premium cigars ^5^. In 2020, cigars became the most popular combustible tobacco product among middle and high school students ^6^. Between 2008 and 2019, the number of imported premium cigars into the US has continuously increased ^7^.

Similar to cigarettes, cigars or premium cigars contain nicotine and toxic chemicals, such as polycyclic aromatic hydrocarbons (PAH) and volatiles. Many studies have shown that cigar smoking was associated with different health risks, including heart diseases, respiratory diseases, and all-cause mortality ^8,9^. It is estimated that around 9,000 deaths per year among adults aged over 35 years in the US might result from cigar smoking ^10^. While there is no direct evidence evaluating the health risks of premium cigars, considering similar chemical constituents between premium cigars and other combustible tobacco products, premium cigars might be associated with some health outcomes, such as respiratory diseases.

Premium cigars are usually promoted by tobacco companies as “handmade” or “premium”, which are normally associated with being better quality and less harmful ^11-13^. While many studies examined public perceptions regarding cigars ^14-18^, studies on public perceptions and discussions of premium cigars are limited ^19,20^. As one of the most popular social media platforms with over 430 million monthly active users, Reddit provides users an ideal venue to share their experiences and perceptions on commonly interested products and topics. Using Reddit data, several previous studies examined public perceptions and discussions of other tobacco products, such as electronic cigarettes ^21,22^.

In this study, we aim to understand public perceptions and discussions of premium cigars on Reddit. We first showed the longitudinal trend for mentions of premium cigars on Reddit from 2019 to 2021. By hand-coding Reddit posts related to premium cigars, we examined public perceptions and discussions of premium cigars. Our findings present valuable evidence on how the public perceives premium cigars on social media, which will provide policymakers with some useful guidance for future regulations on premium cigars to protect public health.

## METHODS

### Data collection

Reddit data between July 2019 and June 2021 were downloaded from the Reddit Archive (http://files.pushshift.io/reddit/comments/). Reddit posts related to Premium cigars were extracted by using two groups of keywords: 1) “cigar” or “cigars” or “cigarillos” or “cigarillo”, and 2) “premium” or “handmade”. In total, 2,238 Reddit posts have been identified.

### Content analysis

To develop the initial codebook, 300 Reddit posts have been randomly sampled from 2,238 posts to identify themes. Two coders manually examined these 300 posts independently. Depending on whether the post directly related to premium cigars, the posts were classified as either relevant or irrelevant. Posts were considered “relevant” if the content explicitly discussed any aspect of premium cigars, either by itself or in comparison to other combustible tobacco products. Posts were deemed “irrelevant” if they simply discussed regular cigars or cigarillos with no mention of or reference to premium cigars. For those posts related to premium cigars, they were summarized into different topics based on their content. Considering the relatively long text for Reddit posts, each post could be assigned to multiple topics. The agreement rate between the two individual coders is 86.44%. Any coding discrepancy between the two coders was resolved by a group of four members. The codebook was developed from these 300 example posts, which was applied to the rest of the Reddit posts. The remaining Reddit posts were split and hand-coded by the two coders.

In total, there were six main topics for “premium cigars”-related Reddit posts, including “Information sharing”, “Affordability of premium cigars”, “User experiences”, “Health related to premium cigars”, “Legal discussion on premium cigars”, and “Mentions of premium cigars”. The topic “Information sharing” has five subtopics, including “Giving suggestions”, “Asking questions”, “Providing reviews”, “Sharing perceptions”, and “Commentary”. The topic “User experiences” includes subtopics “Taste of premium cigars” and “Personal testimony”. The topic “Legal discussion on premium cigars” includes two subtopics “Teenage use” and “Legal regulations on premium cigars”. The topic “Health related to premium cigars” has three subtopics, including “Comparison with cigarettes”, “Addiction and nicotine”, and “Potential health risks”. Appendix Table 1 provided detailed explanation of each topic and subtopic, as well as some Reddit post examples.

## RESULTS

### Longitudinal trends of Reddit posts mentioning premium cigars

Among 2,238 Reddit posts based on the keyword search between July 2019 and June 2021, there were 1,626 posts directly related to premium cigars. The longitudinal trend on the number of posts per month showed that while the number of posts mentioning premium cigars per month remains at a low level (40 – 100 posts each month), the mentions of premium cigars on Reddit greatly increased starting from June 2020, and slowly decreased until March 2021 with an increasing trend afterward (Figure 1).

**Figure 1.**
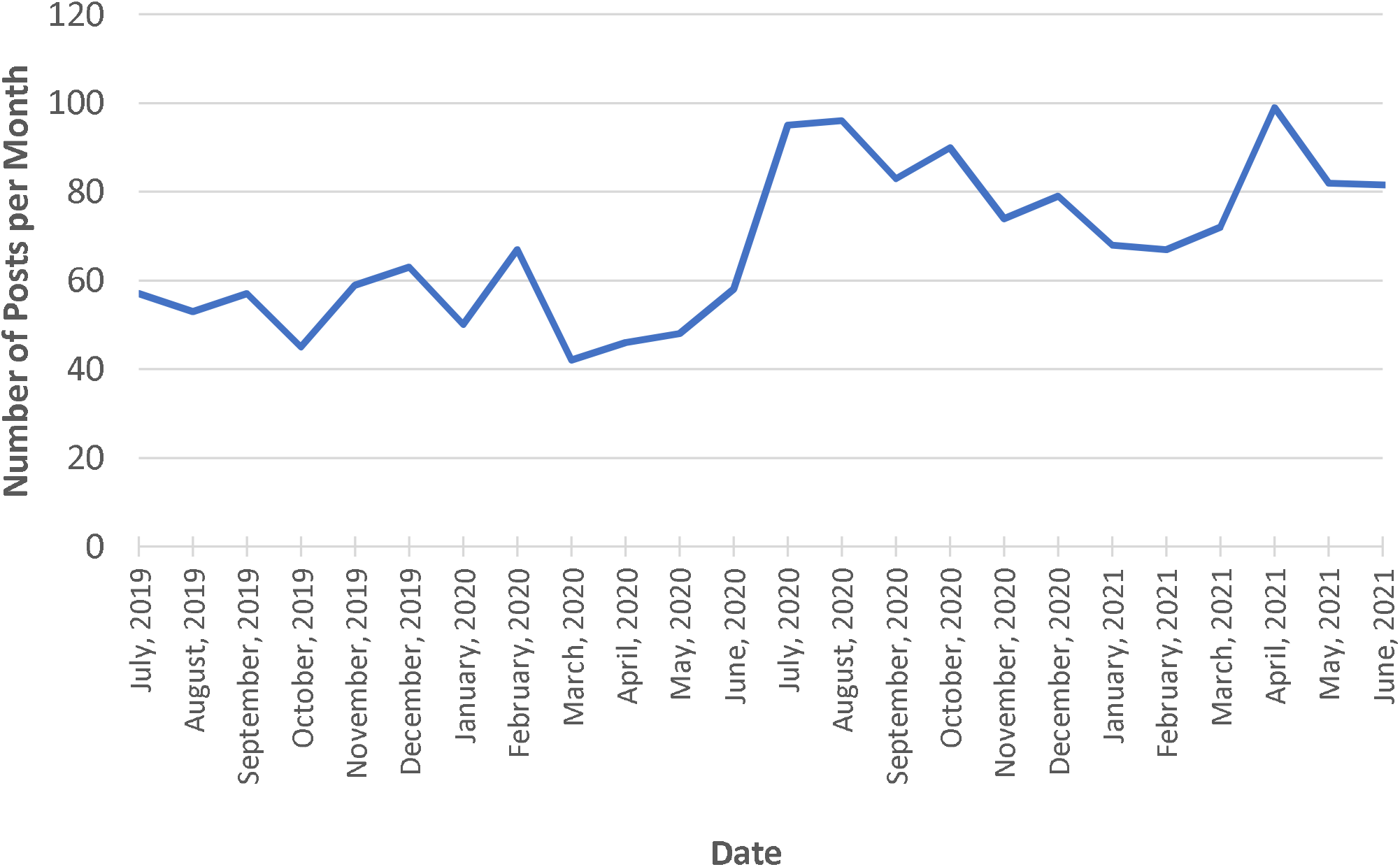
Number of premium cigars-related posts on Reddit over time.

### Topics and subtopics related to premium cigars on Reddit

Through hand-coding, Reddit posts related to premium cigars were grouped into different topics and subtopics depending on their content. As shown in Table 1, the topic “Information sharing” was the dominant (1232 posts/1626 = 75.72%), followed by “User experiences” (442 posts, 27.17%), “Affordability of premium cigars” (309 posts, 18.99%), “Legal discussion on premium cigars” (128 posts, 7.87%), “Health related to premium cigars” (111 posts, 6.82%), and “Mentions of premium cigars” (256 posts, 15.73%). Under the topic “Information sharing”, the subtopic “Sharing perceptions” was the dominant (696 posts, 56.49%), followed by “Giving suggestions” (308 posts, 25%), “Commentary” (251 posts, 20.29%), “Asking questions” (144 posts, 11.69%), and “Providing reviews” (56 posts, 4.55%). Under the topic “User experience”, there were 154 posts (34.84%) about the taste of premium cigars and 288 posts (65.16%) about personal testimony. With the topic of “Health related to premium cigars”, the subtopic “Comparison with cigarettes” has 56 posts (50.45%), and the subtopic “Addiction and nicotine” has 34 posts (30.63%). There were 28 posts (21.88%) about “Teenage use” and 100 posts (78.13%) about “Legal regulations on premium cigars” within the topic “Legal discussion on premium cigars”.

**Table 1.** Topics and subtopics related to premium cigars on Reddit.

| Topics (Number of posts, %) | Subtopics | Number of posts (%) |
| --- | --- | --- |
| Information sharing (1232, 75.72%) |  |  |
|  | Giving suggestions | 308 (25%) |
|  | Asking questions | 144 (11.69%) |
|  | Providing reviews | 56 (4.55%) |
|  | Sharing perceptions | 696 (56.49%) |
|  | Commentary | 251 (20.29%) |
| User experiences (442, 27.17%) |  |  |
|  | Taste of premium cigars | 154 (34.84%) |
|  | Personal testimony | 288 (65.16%) |
| Affordability of premium cigars (309, 18.99%) |  |  |
| Legal discussion on premium cigars (128, 7.87%) |  |  |
|  | Teenage use | (28, 21.88%) |
|  | Legal regulations on premium cigars | (100, 78.13%) |
| Health related to premium cigars (111, 6.82%) |  |  |
|  | Comparison with cigarettes | 56 (50.45%) |
|  | Addiction and nicotine | 34 (30.63%) |
|  | Potential health risks | 37 (33.33%) |
| Mentions of premium cigars (256, 15.73%) |  |  |

## DISCUSSION

In this study, we examined public perceptions and discussions related to premium cigars between 2019 and 2021 using Reddit data. We showed that the mentions of premium cigars on Reddit increased starting from June 2020 and slightly decreased afterward but increased again in April 2021. Our content analysis identified six major topics related to premium cigars, with “Information sharing” as the dominant topic, followed by “User experiences” and “Affordability of premium cigars”. Within the topic “Information sharing”, the subtopic “Sharing perceptions” was the most dominant.

Reddit, as a popular social media platform, is often meant for users to share personal opinions and user experiences on commonly interested products or topics, such as the opinions and user experiences of different e-cigarette flavors ^23 21^. In this study, we showed that about 75% of Reddit posts mentioning premium cigars were sharing information about premium cigars, such as asking questions, giving advice related to premium cigars, and especially sharing opinions about premium cigars. Together with the increasing trend of premium cigars mentioned on Reddit, our results demonstrated that more and more users were curious or interested in premium cigars. In addition, about 27% of Reddit posts were about sharing user experiences (such as how premium cigars taste), suggesting that social media, especially Reddit, has become a popular place for users to share their personal experiences with premium cigars or even other tobacco products, which might affect the prevalence of premium cigar use.

It was reported before that premium cigar users perceived cigars, especially premium cigars, as less harmful and addictive than cigarettes ^19 24 18^. In this study, we showed that many Reddit users perceived premium cigars as less harmful and less addictive compared to cigarettes, for example, “Premium cigars are containing only tobacco. Now there is effectively less chemicals… at least no added chemicals. But the tobacco when burning releases some chemicals that can be bad for your health” and “Premium cigars, when smoked in moderation and not inhaled, are not very addictive. If you abuse it, that’s on you”. Public perceptions describing premium cigars as less harmful and addictive on social media might contribute to the growing prevalence of premium cigar use. These misinformation about premium cigars shared on Reddit, such as less harmful and addictive, can mislead Reddit users, which need to be further regulated.

There are several limitations to this study. First, the number of Reddit posts related to premium cigars collected in this study is relatively small, which might limit more robust analysis and concrete conclusions. Second, while Reddit is one of the most popular social media platforms, Reddit users do not represent the whole population, which might introduce some biases into our findings. According to user distribution statistics collected as of March 2021, about 49% of Reddit users aged 29 or younger and 51% aged 30 or older ^25^, which are slightly different from the US population. Third, since Reddit does not provide the demographics (such as age, gender, or race/ethnicity) and geolocation of users, we are not able to compare the differences in the perceptions and discussions of premium cigars among different demographics of Reddit users who have posted premium cigar-related contents. In addition, some Reddit posts may come from non-US countries considering about a half of Reddit users are from the US. While our results may not fully represent what happened in the US, it still can provide useful information for US tobacco regulation. Lastly, some Reddit posts related to premium cigars might be missed in our study due to the limited number of keywords included in the keyword searching, which might affect our results.

Our study, for the first time, examined public perceptions and discussions of premium cigars on social media using Reddit data. We showed the top topics about premium cigars that have been actively discussed on social media. While our study might be relatively preliminary, our results do provide some important information about premium cigars, for example, users like to share their perceptions and user experiences with premium cigars, and misperception of premium cigars as less harmful and less addictive than cigarettes. These findings might be valuable for future tobacco regulatory policies on premium cigars to protect public health, such as developing campaigns to counter misinformation about the harms of premium cigars use on social media.

## Data Availability

All data produced in the present study are available upon reasonable request to the authors

## Funding

This study was supported by the WNY Center for Research on Flavored Tobacco Products (CRoFT) under cooperative agreement U54CA228110 from the National Cancer Institute of the National Institutes of Health and the U.S. Food and Drug Administration.

## Contributors

ZX, DL: conceived and designed the study. ZX, SL, EX: analyzed the data. ZX, SL, EX and DL: assisted with interpretation of analyses and wrote and edited the manuscript.

## Disclaimer

The content is solely the responsibility of the authors and does not necessarily represent the official views of the NIH or the FDA.

## Declaration of Interests

None declared.

## Data Availability

The data will be shared upon reasonable request to the corresponding author.

## Notes

### Competing Interest Statement

The authors have declared no competing interest.

### Funding Statement

This study was funded by the WNY Center for Research on Flavored Tobacco Products (CRoFT) under cooperative agreement U54CA228110 from the National Cancer Institute of the National Institutes of Health and the U.S. Food and Drug Administration.

